# Association of serum adiponectin and leptin levels with inner retinal thickness among individuals with or without elevated HbA1c

**DOI:** 10.1101/2024.07.01.24309679

**Authors:** Neslihan D Koseoglu, Jiangxia Wang, Frederick Anokye-Danso, Jose Amezcua Moreno, Eumee Cha, Franklin Fuchs, Jacqueline Teed, Jianing Yao, Yan Zhang, Rexford S Ahima, Mira M Sachdeva

**Author notes:** ***Corresponding author***: Mira M Sachdeva, MD, PhD Wilmer Eye Institute Johns Hopkins University School of Medicine 600 N. Wolfe Street, Maumenee 748 Baltimore, MD 21287 **Email**.

## Abstract

Inner retinal thinning precedes clinical evidence of retinopathy in prediabetes and diabetes mellitus (DM), and may contribute to retinopathy development and progression. Serum levels of the adipokines leptin and adiponectin are inversely related in the setting of impaired glucose homeostasis, but their potential association with inner retinal thickness is unknown. In this prospective study, both eyes from 24 individuals with prediabetes or type 2 DM (glycated hemoglobin [HbA1c] ≥5.7) and 16 controls (HbA1c<5.7) underwent spectral-domain optical coherence tomography imaging of the macula, and thickness of the nerve fiber layer (NFL) and ganglion cell layer-inner plexiform layer (GCL-IPL) was analyzed in each subfield of the Early Treatment Diabetic Retinopathy Study grid. Serum samples were collected and metabolic factors, including adiponectin and leptin, were measured. Adjusted regression analyses revealed inverse associations of these adipokines with NFL thickness that did not differ between prediabetes/DM and controls, but differential positive associations of adiponectin with GCL-IPL thickness only in the prediabetes/DM group. The results of our pilot study suggest opposing roles for adiponectin and leptin in the retina, similar to their relationship in systemic disease, and suggest that serum adiponectin may represent a potential clinical biomarker for inner retinal thickness in patients with elevated HbA1c.

## Introduction

Diabetic retinopathy (DR) is one of the most common microvascular complications of diabetes mellitus (DM) and the leading cause of preventable blindness in the working age population despite currently-available treatments, suggesting a need for improved risk stratification and earlier intervention.^1,2^ Recent estimates indicate that approximately 462 million individuals are affected by type 2 DM worldwide, and that the prevalence continues to increase.^3^

The link between elevated serum glucose levels or glycated hemoglobin (HbA1c) and DR has been well-established by several studies including the UK prospective diabetes study (UKPDS).^4^ Prediabetes, defined as HbA1c of 5.7%-6.4%, represents an early stage of glucose dysregulation not reaching the diagnostic threshold for DM (HbA1c>6.4%), that increases risk of type 2 DM and itself can be associated with retinopathy.^5,6^

In addition to chronic hyperglycemia, as indicated by HbA1c levels, several other modifiable metabolic measures including systolic blood pressure (SBP), body mass index (BMI), and dyslipidemia, have been recognized as risk factors for retinopathy.^7,8^ Obesity, defined as BMI of ≥30 kg/m^2^, is associated with altered secretion of adipokines from adipose tissue, significantly influencing the development of DM complications.^9,10^ Serum levels of adiponectin, an adipokine with insulin sensitizing, anti-oxidative, and anti-inflammatory properties, are lower in individuals with obesity, prediabetes, and type 2 DM.^11–13^ Conversely, leptin increases with adiposity and possesses pro-inflammatory characteristics.^10,14^

While DR remains defined and staged based on its retinal microvascular findings, increasing evidence indicates that neurodegenerative changes including thinning of the inner retina precede clinically evident DR, and may even contribute to DR development and progression.^15–20^ Although there are several studies reporting the relationship with adipokines and DR, their potential association with early diabetic retinal neurodegeneration is unknown.^21–25^ In this study, we performed a topographical quantitative assessment of the inner retinal thickness of the macula in individuals with prediabetes or type 2 DM and age/sex-matched controls, and assessed the association of inner retinal thickness with clinically relevant metabolic factors, including adiponectin and leptin.

## Results

A total of 24 individuals with prediabetes or type 2 DM (HbA1c≥5.7%) and 16 controls (HbA1c<5.7%) were included in this study. Age, sex, and metabolic parameters were similar between groups, with the exception of HbA1c and cholesterol **(Table 1)**. Mean nerve fiber layer (NFL) and ganglion cell layer-inner plexiform layer (GCL-IPL) thickness in each Early Treatment Diabetic Retinopathy Study (ETDRS) subfield of the macula was also not different between the groups, potentially due to the small sample size **(Table 2).**

**Table 1:** Baseline characteristics of participants in control and prediabetes/type 2 DM groups.

|  |  | **Control**<br>**(n=16)** | **Prediabetes/type2 DM**<br>**(n=24)** | **p-value** |
| --- | --- | --- | --- | --- |
| **Age (years)<sup>\*</sup>** |  | 70.7 (6.3) | 71.9 (6.2) | 0.56 |
| **Gender<sup>§</sup>** | Female | 9 (56%) | 13 (54%) | 0.90 |
|  | Male | 7 (44%) | 11 (46%) |  |
| **HbA1c (%)<sup>\*</sup>** |  | 5.3 (0.3) | 6.9 (1.3) | **<0.001** |
| **Adiponectin (µg/mL)<sup>\*</sup>** |  | 11.6 (8.6) | 11.0 (9.2) | 0.85 |
| **Leptin (ng/mL)<sup>\*</sup>** |  | 26.8 (21.9) | 24.0 (11.2) | 0.59 |
| **RAGE (ng/mL)<sup>\*</sup>** |  | 11.5 (3.0) | 13.4 (7.1) | 0.34 |
| **NEFA (mM)<sup>\*</sup>** |  | 0.9 (0.2) | 1.0 (0.3) | 0.39 |
| **SBP (mmHg)<sup>\*</sup>** |  | 151.6 (25.4) | 142.9 (20.1) | 0.23 |
| **Cholesterol (mg/dL)<sup>\*</sup>** |  | 224.9 (40.5) | 195.6 (26.7) | **0.009** |
| **Ketone (mM)<sup>\*</sup>** |  | 0.2 (0.4) | 0.1 (0.1) | 0.11 |
| **BUN/Cr<sup>\*</sup>** |  | 15.8 (3.8) | 17.9 (4.2) | 0.13 |

<sup>\*</sup>For continuous variables, means are presented with standard deviations in parentheses, and compared with t-tests
<sup>§</sup>For categorical variables, means are presented with percentage in parentheses, and compared with chi-squared tests
Abbreviations: HbA1c, glycated hemoglobin; RAGE, receptor for advanced glycation end products; NEFA, non-esterified fatty acids; SBP, systolic blood pressure; BUN, blood urea nitrogen; Cr, creatinine

**Table 2:** Comparison of mean NFL and GCL-IPL thickness values between prediabetes/type 2DM and control groups.

| EDTRS subfield | Control<br>(n=32 eyes) | Prediabetes/type 2DM<br>(n=48 eyes) | Total<br>(n=80 eyes) | p-value |
| --- | --- | --- | --- | --- |
| **NFL thickness ($\mu\text{m}$)** |  |  |  |  |
| **CSF** | 13.3 (2.3) | 12.6 (3.9) | 12.9 (3.4) | 0.40 |
| **SI** | 24.1 (3.6) | 23.2 (2.8) | 23.5 (3.1) | 0.21 |
| **SO** | 38.4 (6.9) | 37.2 (4.0) | 37.6 (5.3) | 0.38 |
| **NI** | 21.4 (3.5) | 20.8 (4.8) | 21.0 (4.3) | 0.50 |
| **NO** | 49.0 (7.1) | 48.1 (8.3) | 48.5 (7.8) | 0.56 |
| **II** | 24.1 (3.8) | 23.9 (4.4) | 24.0 (4.2) | 0.68 |
| **IO** | 38.1 (6.8) | 38.0 (6.8) | 38.1 (6.8) | 0.86 |
| **TI** | 17.2 (2.1) | 17.5 (2.1) | 17.4 (2.1) | 0.60 |
| **TO** | 19.2 (2.0) | 19.5 (2.3) | 19.4 (2.1) | 0.75 |
| **GCL-IPL thickness ($\mu\text{m}$)** |  |  |  |  |
| **CSF** | 39.4 (9.5) | 35.8 (10.9) | 37.2 (10.4) | 0.13 |
| **SI** | 87.0 (8.4) | 88.0 (7.9) | 87.6 (8.0) | 0.98 |
| **SO** | 56.5 (5.4) | 59.2 (5.6) | 58.1 (5.6) | 0.15 |
| **NI** | 88.3 (9.2) | 87.2 (8.5) | 87.6 (8.7) | 0.49 |
| **NO** | 59.8 (7.1) | 60.8 (5.8) | 60.4 (6.3) | 0.69 |
| **II** | 86.8 (9.7) | 86.0 (8.3) | 86.3 (8.8) | 0.57 |
| **IO** | 57.6 (6.0) | 57.1 (6.1) | 57.3 (6.0) | 0.72 |
| **TI** | 84.1 (10.2) | 81.9 (8.8) | 82.8 (9.4) | 0.22 |
| **TO** | 62.8 (5.2) | 64.2 (6.4) | 63.7 (6.0) | 0.48 |

Data are presented as mean thickness with standard deviation in parentheses.
Abbreviations: NFL, nerve fiber layer; GCL-IPL, ganglion cell layer-inner plexiform layer; CSF, central subfield; SI, superior inner; SO, superior outer; NI, nasal inner; NO, nasal outer; II, inferior inner; IO, inferior outer; TI, temporal inner; TO, temporal outer

When evaluating the eyes of all participants together, we observed significant positive associations between serum adiponectin levels and NFL thickness in the nasal inner (NI) (0.249 [CI: 0.094, 0.392], p= 0.011), nasal outer (NO) (0.480 [CI: 0.235, 0.712], p=0.003), temporal inner (TI) (0.114 [CI: 0.50, 0.174, p=0.005) and temporal outer (TO) subfields (0.091 [CI:0.022, 0.154], p=0.032), while there were no statistically significant associations between leptin levels and NFL thickness **(Table 3)**.

**Table 3.** Adjusted regression analyses of associations of serum adiponectin and leptin levels with NFL thickness across all ETDRS regions among all participants.

| ETDRS subfield | Adiponectin | p value | Leptin | p value |
| --- | --- | --- | --- | --- |
| **CSF** | 0.104 (0.004-0.205) | 0.095 | 0.012 (-0.043-0.070) | 0.721 |
| **SI** | 0.114 (0.014-0.201) | 0.059 | -0.048 (-0.098-0.007) | 0.158 |
| **SO** | 0.072 (-0.092-0.233) | 0.462 | -0.083 (-0.175-0.010) | 0.143 |
| **NI** | 0.249 (0.094-0.392) | **0.011\*** | 0.012 (-0.063-0.095) | 0.805 |
| **NO** | 0.480 (0.235-0.712) | **0.003\*** | -0.041 (-0.160-0.092) | 0.599 |
| **II** | 0.155 (0.003-0.302) | 0.095 | -0.044 (-0.128-0.042) | 0.396 |
| **IO** | 0.244 (0.050-0.174) | 0.071 | -0.074 (-0.195-0.052) | 0.328 |
| **TI** | 0.114 (0.50-0.174) | **0.005\*** | 0.015 (-0.019-0.051) | 0.476 |
| **TO** | 0.091 (0.022-0.154) | **0.032\*** | 0.007 (-0.029-0.046) | 0.771 |

Values reported as regression coefficient (95% confidence interval).
Asterisk indicates statistical significance.
Abbreviations: NFL, nerve fiber layer; CSF, central subfield; SI, superior inner; SO, superior outer; NI, nasal inner; NO, nasal outer; II, inferior inner; IO, inferior outer; TI, temporal inner; TO, temporal outer

For GCL-IPL thickness, we observed a negative association with leptin across all ETDRS subfields that only reached statistical significance in the superior inner (SI) region (-0.197 [CI: - 0.355, -0.038], p=0.047). Conversely, adiponectin exhibited a significant positive association with GCL-IPL thickness solely in the TO region (0.274 [CI: 0.091, 0.457], p=0.018) **(Table 4)**. However, due to the large number of variables included in our analyses, these associations did not remain significant after multiple comparison adjustment.

**Table 4.** Adjusted regression analyses of associations of serum adiponectin and leptin levels with GCL-IPL thickness across all ETDRS regions among all participants.

| **ETDRS subfield** | **Adiponectin** | **p value** | **Leptin** | **p value** |
| --- | --- | --- | --- | --- |
| **CSF** | 0.203 (-0.168-0.568) | 0.364 | -0.059 (-0.264-0.153) | 0.641 |
| **SI** | 0.114 (-0.162-0.394) | 0.498 | -0.197 (-0.355- -0.038) | **0.047\*** |
| **SO** | 0.194 (0.027-0.366) | 0.069 | -0.085 (-0.812- -0.010) | 0.155 |
| **NI** | 0.017 (-0.303-0.338) | 0.932 | -0.155 (-0.321-0.017) | 0.144 |
| **NO** | -0.050 (-0.288-0.191) | 0.730 | -0.143 (-0.270- -0.017) | 0.070 |
| **II** | 0.066 (-0.238-0.365) | 0.719 | -0.179 (-0.348- -0.005) | 0.094 |
| **IO** | 0.158 (-0.058-0.375) | 0.231 | -0.026 (-0.148-0.098) | 0.727 |
| **TI** | 0.220 (-0.096-0.532) | 0.253 | -0.206 (-0.384- -0.027) | 0.064 |
| **TO** | 0.274 (0.091-0.457) | **0.018\*** | -0.074 (-0.179-0.029) | 0.239 |

Values reported as regression coefficient (95% confidence interval).
Asterisk indicates statistical significance.
Abbreviations: GCL-IPL, ganglion cell layer-inner plexiform layer; CSF, central subfield; SI, superior inner; SO, superior outer; NI, nasal inner; NO, nasal outer; II, inferior inner; IO, inferior outer; TI, temporal inner; TO, temporal outer

Interestingly, regression analyses for the groups separately revealed overall similar positive associations with adiponectin and negative associations with leptin for both NFL and GCL-IPL thickness. Regarding NFL thickness, significant positive associations with adiponectin levels were observed in the prediabetes/type 2DM group in the central subfield (CSF) (0.152 [CI: 0.034, 0.269], p=0.011), NI (0.265 [CI: 0.091, 0.438], p=0.003), NO (0.467 [CI: 0.158, 0.775], p=0.003), inferior inner (II) (0.210 [CI: 0.041, 0.379], p=0.014), and TI (0.103 [CI: 0.032, 0.175], p=0.004) ETDRS subfields, whereas a positive association with NFL was noted only in the SI region for the control group **(Table 5)**. Conversely, leptin showed significant negative associations with NFL in the control group in the SI (-0.076 [CI: -0.129, -0.024], p=0.004), superior outer (SO) (-0.136 [CI: -0.234, -0.039], p=0.006), NO (-0.161 [CI: -0.309, -0.014], p=0.032), and inferior outer (IO) (-0.167 [CI: -0.301, -0.032], p=0.015) ETDRS subfields. There was no association with leptin and NFL thickness in the prediabetes/type 2DM group. Overall, these associations were not significantly different between groups **(Table 5)**.

**Table 5:** Adjusted regression analyses of associations of serum adiponectin and leptin with NFL thickness across all ETDRS regions within control and prediabetes/ type 2DM groups, and differential associations between groups.

| EDTRS subfield | Control |  | Prediabetes/type 2DM |  | Difference between prediabetes/type 2DM versus control |  |
| --- | --- | --- | --- | --- | --- | --- |
| Adiponectin | Coefficient<br>(95% Confidence Interval) | p value | Coefficient<br>(95% Confidence Interval) | p value | Coefficient<br>(95% Confidence Interval) | p value |
| CSF | 0.192<br>(-0.129, 0.167) | 0.799 | 0.152<br>(0.034, 0.269) | **0.011\*** | 0.132<br>(-0.563, 0.321) | 0.169 |
| SI | 0.158<br>(0.015, 0.301) | **0.030\*** | 0.072<br>(-0.041, 0.186) | 0.214 | -0.086<br>(-0.268, 0.096) | 0.356 |
| SO | 0.219<br>(-0.042, 0.480) | 0.100 | -0.031<br>(-0.242, 0.180) | 0.772 | -0.250<br>(-0.586, 0.086) | 0.145 |
| NI | 0.146<br>(-0.046, 0.338) | 0.137 | 0.265<br>(0.091, 0.438) | **0.003\*** | 0.119<br>(-0.139, 0.378) | 0.367 |
| NO | 0.330<br>(-0.010, 0.671) | 0.057 | 0.467<br>(0.158, 0.775) | **0.003\*** | 0.136<br>(-0.323, 0.596) | 0.562 |
| II | 0.007<br>(-0.201, 0.217) | 0.941 | 0.210<br>(0.041, 0.379) | **0.014\*** | 0.202<br>(-0.067, 0.472) | 0.141 |
| IO | 0.286<br>(-0.061, 0.635) | 0.107 | 0.123<br>(-0.157, 0.405) | 0.388 | -0.162<br>(-0.611, 0.285) | 0.477 |
| TI | 0.060<br>(-0.029, 0.150) | 0.187 | 0.103<br>(0.032, 0.175) | **0.004\*** | 0.043<br>(-0.071, 0.158) | 0.462 |
| TO | 0.091<br>(-0.009, 0.192) | 0.075 | 0.053<br>(-0.027, 0.133) | 0.194 | -0.038<br>(-0.167, 0.090) | 0.558 |
| Leptin |  |  |  |  |  |  |
| CSF | -0.013<br>(-0.075, 0.048) | 0.671 | 0.051<br>(-0.045, 0.149) | 0.300 | 0.064<br>(-0.047, 0.177) | 0.260 |
| SI | -0.076<br>(-0.129, -0.024) | **0.004\*** | -0.063<br>(-0.148, 0.021) | 0.145 | 0.013<br>(-0.084, -0.024) | 0.785 |
| SO | -0.136<br>(-0.234, -0.039) | **0.006\*** | -0.051<br>(-0.209, 0.105) | 0.521 | 0.085<br>(-0.095, 0.266) | 0.355 |
| NI | -0.073<br>(-0.156, 0.010) | 0.086 | 0.032<br>(-0.101, 0.167) | 0.632 | 0.105<br>(-0.048, 0.260) | 0.179 |
| NO | -0.161<br>(-0.309, -0.014) | **0.032\*** | -0.129<br>(-0.366, 0.107) | 0.285 | 0.032<br>(-0.240, 0.304) | 0.817 |
| II | -0.083<br>(-0.169, 0.002) | 0.058 | -0.008<br>(-0.144, 0.128) | 0.907 | 0.074<br>(-0.082, 0.232) | 0.351 |
| IO | -0.167<br>(-0.301, -0.032) | **0.015\*** | -0.041<br>(-0.255, 0.172) | 0.706 | 0.126<br>(-0.120, 0.372) | 0.317 |
| TI | -0.007<br>(-0.045, 0.030) | 0.695 | 0.049<br>(-0.011, 0.110) | 0.113 | 0.056<br>(-0.013, 0.126) | 0.111 |
| TO | -0.024<br>(0.064, 0.015) | 0.230 | 0.040<br>(-0.023, 0.104) | 0.215 | 0.064<br>(-0.008, 0.138) | 0.084 |

Asterisk indicates statistical significance. Abbreviations: NFL, nerve fiber layer; CSF, central subfield; SI, superior inner; SO, superior outer; NI, nasal inner; NO, nasal outer; II, inferior inner; IO, inferior outer; TI, temporal inner; TO, temporal outer

Regarding GCL-IPL thickness, adiponectin levels demonstrated positive associations exclusively in the prediabetes/type 2DM group, specifically in the CSF, SO and TO subfields (0.423 [CI: 0.001, 0.843], p= 0.049, 0.335 [CI: 0.120, 0.551], p= 0.002 and 0.387 [CI: 0.166, 0.608], p= 0.001, respectively) **(Table 6).** Interestingly, there was a differential association of adiponectin with GCL-IPL thickness in the prediabetes/type 2DM versus control groups in the SO (p=0.005), NO (p=0.036), and TO (p=0.012) subfields, suggesting that serum adiponectin levels may represent a biomarker for GCL-IPL thickness specifically in the setting of prediabetes/type 2DM. Leptin showed significant negative associations in SI (-0.164 [CI: - 0.324, -0.004], p=0.044) and TI (-0.215 [CI: -0.399, -0.031], p=0.022) subfields in the control group and the SI subfield in the prediabetes/type 2DM group (-0.261 [CI: -0.520, -0.002], p=0.048), with no differential association between groups in any subfield **(Table 6)**. Due to the large number of variables included in our study design, these associations did not remain statistically significant after multiple comparison adjustment.

**Table 6:** Adjusted regression analyses of associations of serum adiponectin and leptin with GCL-IPL thickness across all ETDRS regions within control and prediabetes/ type 2DM groups, and differential associations between groups.

| ETDRS subfield | Control |  | Prediabetes/type 2DM |  | Difference between prediabetes/type 2DM versus control |  |
| --- | --- | --- | --- | --- | --- | --- |
| Adiponectin | Coefficient<br>(95% Confidence Interval) | p value | Coefficient<br>(95% Confidence Interval) | p value | Coefficient<br>(95% Confidence Interval) | p value |
| **CSF** | 0.075<br>(-0.447, 0.597) | 0.778 | 0.423<br>(0.001, 0.843) | **0.049\*** | 0.348<br>(-0.323, 1.019) | 0.310 |
| **SI** | -0.155<br>(-0.582, 0.272) | 0.477 | 0.271<br>(-0.076, 0.619) | 0.126 | 0.426<br>(-0.125, 0.978) | 0.130 |
| **SO** | -0.156<br>(-0.422, 0.110) | 0.251 | 0.335<br>(0.120, 0.551) | **0.002\*\*** | 0.492<br>(0.148, 0.835) | **0.005\*** |
| **NI** | -0.269<br>(-0.691, 0.152) | 0.211 | 0.343<br>(-0.421, 0.729) | 0.081 | 0.613<br>(0.410, 1.185) | **0.036\*** |
| **NO** | -0.220<br>(-0.565, 0.125) | 0.212 | 0.109<br>(-0.208, 0.426) | 0.501 | 0.329<br>(-0.140, 0.798) | 0.170 |
| **II** | -0.233<br>(-0.692, 0.224) | 0.318 | 0.213<br>(-0.155, 0.582) | 0.256 | 0.447<br>(-0.141, 1.036) | 0.137 |
| **IO** | 0.111<br>(-0.199, 0.422) | 0.483 | 0.140<br>(-0.110, 0.391) | 0.272 | 0.029<br>(-0.370, 0.429) | 0.886 |
| **TI** | -0.027<br>(-0.508, 0.453) | 0.911 | 0.369<br>(-0.020, 0.759) | 0.063 | 0.397<br>(-0.222, 1.016) | 0.209 |
| **TO** | -0.065<br>(-0.339, 0.209) | 0.642 | 0.387<br>(0.166, 0.608) | **0.001\*** | 0.452<br>(0.995, 0.805) | **0.012\*** |
| **Leptin** |  |  |  |  |  |  |
| **CSF** | -0.160<br>(-0.370, 0.049) | 0.134 | 0.063<br>(-0.272, 0.399) | 0.711 | 0.224<br>(-0.162, 0.611) | 0.256 |
| **SI** | -0.164<br>(-0.324, -0.004) | **0.044\*** | -0.261<br>(-0.520, -0.002) | **0.048\*** | -0.096<br>(-0.393, 0.200) | 0.524 |
| **SO** | -0.070<br>(-0.187, 0.045) | 0.235 | -0.011<br>(-0.199, 0.176) | 0.904 | 0.059<br>(-0.156, 0.275) | 0.591 |
| **NI** | -0.129<br>(-0.296, 0.037) | 0.129 | -0.219<br>(-0.488, 0.049) | 0.110 | -0.089<br>(-0.398, 0.218) | 0.568 |
| **NO** | -0.094<br>(-0.229, 0.040) | 0.170 | -0.123<br>(-0.340, 0.094) | 0.266 | -0.028<br>(-0.278, 0.220) | 0.820 |
| **II** | -0.160<br>(-0.336, 0.015) | 0.075 | -0.213<br>(-0.493, 0.065) | 0.134 | -0.053<br>(-0.376, 0.269) | 0.745 |
| **IO** | -0.056<br>(-0.181, 0.067) | 0.370 | 0.011<br>(-0.186, 0.210) | 0.907 | 0.068<br>(-0.160, 0.297) | 0.556 |
| **TI** | -0.215<br>(-0.399, -0.031) | **0.022\*** | -0.196<br>(-0.493, 0.099) | 0.194 | 0.018<br>(-0.321, 0.358) | 0.915 |
| **TO** | -0.081<br>(-0.201, 0.039) | 0.187 | -0.055<br>(-0.250, 0.140) | 0.580 | 0.026<br>(-0.197, 0.249) | 0.819 |

Asterisk indicates statistical significance. Abbreviations: GCL-IPL, ganglion cell layer-inner plexiform layer; CSF, central subfield; SI, superior inner; SO, superior outer; NI, nasal inner; NO, nasal outer; II, inferior inner; IO, inferior outer; TI, temporal inner; TO, temporal outer

The associations of the other metabolic factors with NFL and GCL-IPL thickness are shown in Supplementary Tables 1 and 2. Notably, serum receptor for advanced glycation end products (RAGE) and non-esterified fatty acid (NEFA) levels exhibited statistically significant associations only with GCL-IPL thickness. For RAGE, there was a negative association with GCL-IPL thickness in the NO (-1.127 [CI: -2.082, -0.171]) and TO (-0.880 [CI: -1.741, -0.019) subfields within the control group only (p=0.021 and p=0.045, respectively) and a differential association between control and prediabetes/type 2DM groups in the NO subfield (1.079 [CI: 0.066, 2.091], p=0.037). NEFA showed negative associations with NO, II and TO regions in the control group (-15.707 [CI: -29.677, -1.738] p=0.028, -22.617 [CI: -40.895, -4.359] p=0.015 and -12.846 [CI: - 25.434, -0.258] p=0.045, respectively), with no differential association between control and prediabetes/type 2DM in any subfield.

Other metabolic variables, such as SBP, cholesterol, BUN/Cr and ketones displayed sporadic associations with inner retinal thickness measurements across the ETDRS regions for both groups (see **Supplementary Tables 1 and 2).**

## Discussion

Inner retinal thinning, specifically of the GCL-IPL complex and in some reports the NFL, precedes overt DR. ^26,27^ In this pilot study, our aim was to evaluate associations between metabolic factors related to prediabetes/type 2 DM and topographical thickness of these retinal layers, as assessed by OCT. Adiponectin and leptin, two of the major adipokines, have inverse levels in obesity and diabetes, and opposing roles in systemic inflammation and insulin sensitivity.^28^ Overall, our observations revealed positive associations between adiponectin and inner retinal thickness, while associations with leptin were negative.

Adiponectin is the most abundant hormone secreted by adipocytes (i.e. an adipokine), and has potent insulin sensitizing, anti-inflammatory and anti-oxidative actions.^14^ Serum adiponectin levels correlate inversely with obesity and adiposity and exert a protective role in the pathophysiology of DM.^12^ Adiponectin has also been implicated in neurodegeneration and dementia.^29^ However, the role of adiponectin in diabetic eye disease is unclear. Yang et al. found elevated serum and aqueous humor (AH) adiponectin levels in individuals with DR compared with nondiabetic controls, and correlated adiponectin levels with progression of DR.^22^ In a subsequent study, they further observed a significant association between DR severity and total central foveal thickness with concentrations of adiponectin in the AH.^30^ Other reports have presented evidence contradicting a role of adiponectin in DR.^31–33^ To our knowledge, despite multiple studies investigating the association of adiponectin with microvascular complications in the retina, and one study that assessed peripapillary NFL thickness, ours is the first to evaluate its association with inner neuroretinal thickness in the macula.^34^ We observed a positive association of adiponectin with NFL thickness in the nasal and temporal quadrants of the macula among all participants. Similarly, a positive association was also observed between adiponectin and GCL-IPL in the TO subfield. When stratified into prediabetes/type2DM and control groups, we observed that these associations were mostly positive for both NFL and GCL-IPL thickness. Notably, there was a significant difference in these associations between groups for GCL-IPL, with adiponectin levels significantly associated with GCL-IPL thickness only in the prediabetes/type 2DM group. These results suggest that adiponectin may represent a candidate serum biomarker for the early diabetic retinal neurodegeneration that precedes DR.

Leptin is an adipokine known for its role in obesity and regulation of feeding.^35^ Leptin also regulates the neuroendocrine axis and glucose metabolism and has pro-inflammatory actions.^14,35^ Obesity is characterized by hyperleptinemia, leptin resistance, insulin resistance, chronic inflammation, and increased risk of type 2 DM and cardiovascular disease.^36,37^ Increased leptin levels may contribute to the development and progression of retinopathy.^23,24,38–40^ Overall, inflammation associated with increased levels of leptin has been implicated in promoting angiogenesis and oxidative stress exacerbating DR.^23,39,40^ However, some reports have not found a relationship between leptin and DR.^25,38^ In our study, associations between leptin and NFL and GCL-IPL were mainly negative, reaching significance predominantly in the control group and with no significant difference between control and prediabetes/type2DM groups. This result suggests that metabolic associations with leptin and inner retinal thickness are independent of blood glucose levels.

Various other clinical or metabolic factors can also influence the risk of DR development. Increased anaerobic glycolysis, lipid peroxidation, and generation of advanced glycation end-products (AGE) in response to increased glucose levels have been associated with progression of DR.^41^ AGEs are formed through Maillard reaction of a carbonyl group of glucose, lipid or amino acids. Accumulation of AGEs in diabetic patients induces oxidative stress in the retina by interacting with the RAGE receptors on cell membranes. This continuous oxidative damage may contribute to progression to DR through activation of proinflammatory factors and increased expression of vascular endothelial growth factor (VEGF).^42,43^ In DM, reduced insulin action promotes lipolysis and an increase in plasma NEFA levels. The rise in serum NEFA impairs normal lipid metabolism further and results in accumulation of NEFA in insulin-sensitive tissues causing adipose tissue inflammation and contributing to insulin resistance.^44,45^ To date, potential associations between serum RAGE and NEFA levels with inner retinal thickness have not been reported. Our study found a negative association between serum RAGE levels and GCL-IPL thickness in the outer nasal and temporal regions of the macula in the control group, and this association was significantly different than the prediabetes/type 2DM group for NO only. We observed that serum NEFA levels were also negatively associated with NO, II and TO regions only in the control group.

While our study identified associations of serum adipokines with inner retinal thickness, there are limitations. First, this pilot study included a small sample size. Second, the large number of variables (retinal layers, ETDRS subfields, metabolic parameters) included in the regression analyses necessitated correction for multiple comparisons, which reduced the significance of our unadjusted results. Third, due to the small sample size, we considered participants with prediabetes and type 2 DM together rather than separating those with prediabetes from type 2 DM into separate groups. We postulate that comparing these associations between normoglycemic, prediabetes, and type 2 DM groups separately could offer additional insights into potential serum indicators of neuroretinal thickness changes that precede overt diabetes.

Overall, in the prediabetes/type 2DM group, serum adiponectin levels demonstrated positive associations with inner retinal thickness, whereas in control participants leptin was negatively associated, indicating an opposing relationship with neuroretinal thickness similar to their opposing roles in systemic disease. Interestingly, we observed a differential association between adiponectin and GCL-IPL thickness in the prediabetes/type 2DM group compared with controls. The findings of this pilot study suggest that in the setting of impaired glucose homeostasis (elevated HbA1c), serum adiponectin may be a biomarker for inner retinal structural changes relevant to diabetic eye disease.

## Methods

### Subject recruitment

Adults (>18 years of age) with and without type 2 DM were prospectively recruited from the Wilmer Eye Clinics at Johns Hopkins University School of Medicine. Potentially-eligible participants with type 2 DM were identified prior to their clinic visit by review of the electronic medical record (EMR), with the diagnosis of diabetes determined by the American Diabetes Association (ADA) laboratory criteria of fasting serum glucose > 126 mg/dL or glycated hemoglobin (HbA1c) > 6.4% documented within the prior 12 months. Individuals with no documented history of DM in their EMR were considered as potentially-eligible controls. Spectral domain optical coherence tomography (OCT) of the macula was obtained on each participant, and individuals with macular findings such as drusen, epiretinal membrane, and macular edema were excluded. Additional exclusion criteria included history of uveitis, glaucoma, optic neuropathy, high myopia (> 5 diopters by refraction), other retinal pathology, neurodegenerative disease, and history of prior intraocular treatment (including laser or intravitreal injection) or surgery. This study adhered to the tenets of the Declaration of Helsinki and was approved by the Institutional Review Board at Johns Hopkins University School of Medicine. Written informed consent was obtained from each participant.

### Retinal thickness measurement

Spectral-domain OCT images of the macula of each eye were obtained for each participant on the Spectralis instrument (Heidelberg Engineering, Heidelberg, Germany) using the following scan acquisition parameters: dense volume scan (20° × 20°, roughly 6 × 6 mm), 49 B-scans each spaced 120 μm apart, automatic real-time mean of 16, high speed (512 A-scans/B-scan). All images were acquired by the same operator after pupillary dilation with 2.5% phenylephrine and 1% tropicamide. Images were evaluated using the Heidelberg Eye Explorer (Heyex) platform. The automated segmentation tool was used to identify the boundaries of the retinal layers, and scan quality and segmentation accuracy were verified for each individual B scan by two masked graders (*NDK, EC*). Minor manual adjustments to the segmentation were made if needed. Retinal thickness values were obtained for the NFL and GCL-IPL complex in each of the 9 ETDRS subfields, i.e. the central fovea (1mm diameter) and the superior, inferior, nasal, and temporal inner (ring centered on fovea with diameter of 3mm) and outer (ring centered on fovea with diameter of 6mm) subfields. These subfields were indicated as follows: central subfield (CSF), superior inner (SI), superior outer (SO), nasal inner (NI), nasal outer (NO), inferior inner (II), inferior outer (IO), temporal inner (TI) and temporal outer (TO).

### Analysis of metabolic parameters and serum samples

Serum samples were collected, aliquoted, and immediately stored at -80 degrees for further analyses. Adiponectin (Crystal Chem, Catalog #80571), leptin (Crystal Chem, Catalog #80968), and RAGE (Fisher, Catalog #DRG00) were quantitatively analyzed by ELISA, and beta-hydroxybutyric acid (Stanbio, Catalog # 2440-058) and NEFA (Wako NEFA-HR(2), Wako Pure Chemical Industries Ltd., Osaka, Japan) levels assessed by enzymatic colorimetric assay according to manufacturers’ protocols. Values for SBP, HbA1c, BUN, creatinine, and cholesterol on the day of serum collection were extracted from the EMR. As several participants without type 2 DM were noted to have an elevated HbA1c in the prediabetes range, we considered individuals with HbA1c ≥5.7% as the prediabetes/type 2DM group and those <5.7% were considered as controls. All individuals in the control group were confirmed to not be taking anti-hyperglycemic medications.

### Statistical Analyses

Patient demographics and clinical parameters were compared between the control and the prediabetes/type 2 DM groups. Chi-squared tests were used to compare the categorical variables, and the student’s t-tests were used for the continuous variables. To compare the retinal thickness in different ETDRS subfields between the two groups, the linear mixed effects models with a random intercept were used to account for the correlation between measurements from two eyes from the same participant. To investigate the associations between the retinal thickness outcomes and the metabolic parameters, and how they differed between the two groups, similar linear mixed effects models with a random intercept were utilized, including interaction terms between the indicator variable of prediabetes/type 2DM group and the metabolic variables, adjusting for age and sex. The metabolic variables were centered around their sample means. All the analyses were carried out using the statistical software, Stata version 17.0. A p value less than or equal to 0.05 was considered as statistical significance.

## Supporting information

KoseogluEtAl_SupplementaryData

## Data Availability

All data generated or analyzed during this study are included in this manuscript and its supplementary information files.

## Author Contributions

NDK, JW, FAD, JAM, EC, and MMS contributed to data acquisition and retinal image annotation. NDK, JW, EC, FF, JT, JY, YZ, and MMS performed statistical analyses and/or data interpretation. NDK, JW, RSA, and MMS prepared the manuscript. All authors reviewed and approved the final manuscript.

## Acknowledgements

This study was funded by a Doris Duke Foundation Clinical Scientist Development Award (MMS). MMS holds a Wilmer Rising Professorship.

## Additional Information

The authors declare no competing interests.

