## Supplementary material for "Association of serum adiponectin and leptin levels with inner retinal thickness among individuals with or without elevated HbA1c": KoseogluEtAl_SupplementaryData

**Supplementary Tables**

**Supplementary Table 1:** Adjusted regression analyses of associations of SBP and serum HbA1c, cholesterol, BUN/Cr ratio, ketone, RAGE and NEFA levels with NFL thickness across all ETDRS regions within control and prediabetes/type 2DM groups, and differential associations between groups.

| **EDTRS subfield** | **Control** | | **Prediabetes/type 2DM** | | **Difference between prediabetes/type 2DM versus control** | |
|---|---|---|---|---|---|---|
| **SBP** | Coefficient (95% Confidence Interval) | p value | Coefficient (95% Confidence Interval) | p value | Coefficient (95% Confidence Interval) | p value |
| **CSF** | 0.020 (-0.030-0.071) | 0.430 | -0.035 (-0.088-0.018) | 0.200 | -0.055 (-0.128-0.017) | 0.137 |
| **SI** | 0.011 (-0.037-0.060) | 0.658 | -0.026 (-0.079-0.025) | 0.318 | -0.037 (-0.108-0.033) | 0.297 |
| **SO** | 0.010 (-0.078-0.100) | 0.813 | -0.033 (-0.127-0.059) | 0.480 | -0.044 (-0.172-0.083) | 0.496 |
| **NI** | -0.006 (-0.074-0.062) | 0.860 | -0.061 (-0.135-0.126) | 0.104 | -0.055 (-0.154-0.044) | 0.277 |
| **NO** | 0.007 (-0.123-0.137) | 0.916 | -0.037 (-0.176-0.102) | 0.600 | -0.044 (-0.233-0.144) | 0.645 |
| **II** | 0.009 (-0.064-0.084) | 0.795 | -0.025 (-0.102-0.051) | 0.512 | -0.035 (-0.141-0.070) | 0.510 |
| **IO** | 0.033 (-0.087-0.153) | 0.590 | -0.012 (-0.137-0.111) | 0.839 | -0.046 (-0.217-0.125) | 0.599 |
| **TI** | 0.005 (-0.027-0.038) | 0.728 | -0.008 (-0.044-0.026) | 0.622 | -0.014 (-.062-0.033) | 0.546 |
| **TO** | 0.0031 (-0.028-0.035) | 0.847 | -0.039 (-0.073- -0.005) | **0.024*** | -0.042 (-0.089-0.003) | 0.072 |
| **HbA1c** | Coefficient (95% Confidence Interval) | p value | Coefficient (95% Confidence Interval) | p value | Coefficient (95% Confidence Interval) | p value |
| **CSF** | -1.441 (-6.863-3.980) | 0.602 | 0.162 (-0.735-1.059) | 0.723 | 1.603 (-3.907-7.114) | 0.569 |
| **SI** | -0.195 (-5.315-4.924) | 0.940 | -0.012 (-0.884-0.859) | 0.978 | 0.182 (-5.023-5.389) | 0.945 |
| **SO** | -7.257 (-15.851-1.336) | 0.098 | 0.608 (-0.881-2.098) | 0.424 | 7.866 (-0.878-16.610) | 0.078 |
| **NI** | 1.363 (-6.033-8.761) | 0.718 | 0.168 (-1.127-1.464) | 0.799 | -1.195 (-8.727-6.337) | 0.756 |
| **NO** | -2.936 (-16.204-10.331) | 0.664 | -0.295 (-2.639-2.048) | 0.805 | 2.641 (-10.872-16.154) | 0.702 |
| **II** | 2.570 (-4.969-10.109) | 0.504 | 0.074 (-1.205-1.354) | 0.909 | -2.495 (-10.161-5.169) | 0.523 |
| **IO** | -4.902 (-1.871-7.065) | 0.422 | 0.327 (-1.721-2.375) | 0.754 | 5.230 (-6.941-17.402) | 0.400 |
| **TI** | 2.003 (-1.370-5.377) | 0.244 | 0.029 -0.542-0.600) | 0.921 | -1.974 (-5.405-1.455) | 0.259 |
| **TO** | 1.315 (-2.229-4.860) | 0.467 | 0.235 (-0.367-0.837) | 0.444 | -1.080 (-4.684-2.524) | 0.557 |

| Cholesterol | Coefficient (95% Confidence Interval) | p value | Coefficient (95% Confidence Interval) | p value | Coefficient (95% Confidence Interval) | p value |
|---|---|---|---|---|---|---|
| **CSF** | 0.016 (-0.019-0.051) | 0.371 | 0.021 (-.019-0.062 | 0.305 | 0.005 (-0.046-0.057) | 0.841 |
| **SI** | 0.002 (-0.31-0.036) | 0.875 | 0.015 (-0.026-0.056) | 0.473 | 0.012 (-0.039-0.063) | 0.640 |
| **SO** | -0.071 (-0.128-0.015) | 0.013 | -0.014 (-0.082-0.053) | 0.681 | 0.057 (-0.027-0.142) | 0.187 |
| **NI** | 0.008 (-0.041-0.058) | 0.734 | 0.000 (-0.058-0.060) | 0.982 | -0.007 (-0.082-0.066) | 0.834 |
| **NO** | -0.034 (-0.123-0.053) | 0.443 | -0.017 (-0.124-0.088) | 0.743 | 0.016 (-0.115-0.149) | 0.803 |
| **II** | -0.013 (-0.064-0.037) | 0.595 | -0.001 (-0.061-0.058) | 0.967 | 0.012 (-0.062-0.088) | 0.745 |
| **IO** | -0.067 (-0.146-0.011) | 0.094 | -0.003 (-0.096-0.089) | 0.941 | 0.063 (-0.053-0.181) | 0.286 |
| **TI** | 0.008 (-0.013-0.312) | 0.444 | 0.008 (-0.018-0.035) | 0.528 | -0.000 (-0.033-0.033) | 0.995 |
| **TO** | -0.004 (-0.027-0.019) | 0.732 | 0.001 (-0.026-0.030) | 0.912 | -0.005 (-0.029-0.041) | 0.752 |
| **BUN/Cr** | Coefficient (95% Confidence Interval) | p value | Coefficient (95% Confidence Interval) | p value | Coefficient (95% Confidence Interval) | p value |
| **CSF** | -0.104 (-0.457-0.249) | 0.564 | 0.201 (-0.049-0.452) | 0.115 | 0.305 (-0.113-0.724) | 0.153 |
| **SI** | 0.156 (-0.179-0.492) | 0.362 | 0.009 (-0.230-0.248) | 0.940 | -0.147 (-0.545-0.251) | 0.469 |
| **SO** | 1.074 (0.560-1.588) | **0.000*** | -0.056 (-0.425-0.311) | 0.763 | -1.131 (-1.741- -0.521) | **0.000*** |
| **NI** | -0.121 (-0.619-0.375) | 0.631 | 0.192 (-0.158-0.543) | 0.282 | 0.314 (-0.271-0.900) | 0.293 |
| **NO** | 0.181 (-0.726-1.089) | 0.696 | 0.245 (-0.399-0.890) | 0.455 | 0.643 (-1.006-1.135) | 0.906 |
| **II** | -0.319 (-0.833-0.193) | 0.222 | 0.566 (-0.308-0.422) | 0.761 | 0.376 (-0.231-0.984) | 0.225 |
| **IO** | 0.518 (-0.298-1.334) | 0.213 | 0.225 (-0.358-0.808) | 0.449 | -0.293 (-1.259-0.673) | 0.553 |
| **TI** | -0.125 (-0.350-0.100) | 0.277 | 0.035 (-0.124-0.196) | 0.660 | 0.160 (-0.106-0.428) | 0.237 |
| **TO** | 0.001 (-0.240-0.243) | 0.989 | -0.014 (-0.187-0.157) | 0.865 | -0.016 (-0.303-0.270) | 0.910 |
| **Ketone** | Coefficient (95% Confidence Interval) | p value | Coefficient (95% Confidence Interval) | p value | Coefficient (95% Confidence Interval) | p value |
| **CSF** | -3.691 (-7.287- -0.095) | **0.044*** | 3.394 (-16.020-22.809) | 0.732 | 7.096 (-12.769-26.941) | 0.484 |
| **SI** | 0.951 (-2.474-4.378) | 0.586 | -12.654 (-31.225-5.915) | 0.182 | -13.606 (-32.602-5.388) | 0.160 |
| **SO** | -1.225 (-7.584-5.133) | 0.706 | -25.698 (-60.014-8.618) | 0.142 | -24.472 (-59.562-10.617) | 0.172 |
| **NI** | 0.704 (-4.488-5.898) | 0.790 | -3.581 (-32.622-25.459) | 0.809 | -4.286 (-33.908-25.335) | 0.777 |
| **NO** | 0.0359 (-8.883-9.602) | 0.939 | -33.733 (-85.409-17.941) | 0.201 | -34.093 (-86.796-18.609) | 0.205 |

| | | | | | | |
|---|---|---|---|---|---|---|
| **II** | 1.025 (-4.364-6.415) | 0.709 | -10.483 (-39.452-18.485) | 0.478 | -11.509 (-41.131-18.113) | 0.446 |
| **IO** | -0.912 (-9.386-7.560) | 0.833 | -38.117 (-83.665-7.430) | 0.101 | -37.204 (-83.778-9.368) | 0.117 |
| **TI** | 0.051 (-2.311-2.413) | 0.966 | -2.071 (-14.880-10.738) | 0.751 | -2.122 (-15.224-10.980) | 0.751 |
| **TO** | 0.009 (-2.414-2.433) | 0.994 | -10.447 (-23.582-2.688) | 0.119 | -10.456 (-23.892-2.979) | 0.127 |
| **RAGE** | Coefficient (95% Confidence Interval) | p value | Coefficient (95% Confidence Interval) | p value | Coefficient (95% Confidence Interval) | p value |
| **CSF** | 0.099 (-0.369-0.568) | 0.677 | -0.071 (-0.224-0.081) | 0.359 | -0.171 (-0.666-0.324) | 0.498 |
| **SI** | 0.039 (-0.406-0.486) | 0.861 | 0.025 (-0.122-0.174) | 0.731 | -0.013 (-0.486-0.458) | 0.954 |
| **SO** | -0.432 (-1.207-0.341) | 0.273 | 0.012 (-0.249-0.274) | 0.927 | 0.445 (-0.376-1.266) | 0.288 |
| **NI** | 0.339 (-0.296-0.975) | 0.295 | -0.011 (-0.223-0.199) | 0.912 | -0.351 (-1.024-0.321) | 0.306 |
| **NO** | 0.267 (-0.884-1.420) | 0.649 | 0.773 (-0.309-0.464) | 0.695 | -0.190 (-1.411-1.030) | 0.760 |
| **II** | 0.251 (-0.401-0.903) | 0.450 | -0.064 (-0.282-0.154) | 0.565 | -0.315 (-1.006-0.375) | 0.371 |
| **IO** | 0.159 (-0.894-1.213) | 0.767 | 0.025 (-0.329-0.380) | 0.888 | -0.134 (-1.251-0.983) | 0.814 |
| **TI** | 0.097 (-0.192-0.388) | 0.510 | -0.049 (-0.145-0.045) | 0.306 | -0.147 (-0.455-0.159) | 0.346 |
| **TO** | 0.069 (-0.237-0.376) | 0.658 | -0.053 (-0.155-0.048) | 0.302 | -0.123 (-0.448-0.202) | 0.458 |
| **NEFA** | Coefficient (95% Confidence Interval) | p value | Coefficient (95% Confidence Interval) | p value | Coefficient (95% Confidence Interval) | p value |
| **CSF** | -2.224 (-8.826-4.377) | 0.509 | -0.621 (-4.707-3.463) | 0.765 | 1.602 (-6.213-9.418) | 0.688 |
| **SI** | -5.292 (-11.308-0.722) | 0.085 | -2.110 (-5.840-1.618) | 0.267 | 3.182 (-3.944-10.308) | 0.381 |
| **SO** | -9.293 (-20.083-1.496) | 0.091 | -4.347 (-11.103-2.408) | 0.207 | 4.945 (-7.850-17.741) | 0.449 |
| **NI** | -2.534 (-11.568-6.500) | 0.582 | -2.115 (-7.725-3.494) | 0.460 | 0.418 (-10.256-11.094) | 0.939 |
| **NO** | -7.861 (-23.883-8.159) | 0.336 | -6.679 (-16.664-3.304) | 0.190 | 1.182 (-17.759-20.123) | 0.903 |
| **II** | -3.551 (-12.926-5.822) | 0.458 | -1.519 (-7.362-4.324) | 0.610 | 2.032 (-9.068-13.134) | 0.720 |
| **IO** | -8.385 (-22.901-6.129) | 0.258 | -7.535 (-16.602-1.532) | 0.103 | 0.850 (-16.345-18.046) | 0.923 |
| **TI** | -0.575 (-4.731-3.581) | 0.786 | 0.622 (-1.950-3.195) | 0.635 | 1.197 (-3.725-6.120) | 0.633 |
| **TO** | -1.923 (-6.230-2.383) | 0.381 | -1.526 (-4.196-1.144) | 0.263 | 0.397 (-4.705-5.499) | 0.879 |

Values reported as regression coefficient (95% confidence interval).
Asterisk indicates statistical significance.
Abbreviations: NFL, nerve fiber layer; SI, superior inner; SO, superior outer; NI, nasal inner; NO, nasal outer; II, inferior inner; IO, inferior outer; TI, temporal inner; TO, temporal outer; SBP, systolic blood pressure; HbA1c, glycated hemoglobin; BUN/Cr, blood urea nitrogen/creatinine; RAGE, receptor for advanced glycation end products; NEFA, non-esterified fatty acids

**Supplementary Table 2:** Adjusted regression analyses of associations of SBP and serum HbA1c, cholesterol, BUN/Cr ratio, ketone, RAGE and NEFA levels with GCL-IPL thickness across all ETDRS regions within control and prediabetes/type 2DM groups, and differential associations between groups.

| EDTRS subfield | Control | | Prediabetes/type 2DM | | Difference between prediabetes/type 2DM versus control | |
|---|---|---|---|---|---|---|
| **SBP** | Coefficient (95% Confidence Interval) | p value | Coefficient (95% Confidence Interval) | p value | Coefficient (95% Confidence Interval) | p value |
| **CSF** | -0.022 (-0.200-0.156) | 0.808 | -0.124 (-0.308-0.597) | 0.185 | -0.102 (-0.356-0.151) | 0.429 |
| **SI** | 0.020 (-0.127-0.168) | 0.788 | 0.034 (-0.118-0.188) | 0.658 | 0.014 (-0.196-0.225) | 0.894 |
| **SO** | 0.026 (-0.073-0.126) | 0.606 | -0.030 (-0.134-0.073) | 0.567 | -0.056 (-0.199-0.086) | 0.437 |
| **NI** | 0.038 (-0.109-0.187) | 0.606 | -0.024 (-0.181-0.132) | 0.760 | -0.063 (-0.276-0.149) | 0.560 |
| **NO** | 0.056 (-0.060-0.173) | 0.344 | -0.035 (-0.157-0.086) | 0.569 | -0.091 (-0.259-0.086) | 0.282 |
| **II** | 0.071 (-0.084-0.228) | 0.366 | 0.017 (-0.143-0.178) | 0.832 | -0.054 (-0.276-0.167) | 0.631 |
| **IO** | 0.038 (-0.065-0.141) | 0.469 | -0.074 (-0.180-0.032) | 0.171 | -0.112 (-0.258-0.034) | 0.134 |
| **TI** | 0.071 (-0.095-0.237) | 0.403 | -0.023 (-0.196-0.149) | 0.789 | -0.094 (-0.332-0.143) | 0.435 |
| **TO** | 0.581 (-0.043-0.159) | 0.261 | -0.065 (-0.171-0.040) | 0.223 | -0.124 (-0.269-0.021) | 0.094 |
| **HbA1c** | Coefficient (95% Confidence Interval) | p value | Coefficient (95% Confidence Interval) | p value | Coefficient (95% Confidence Interval) | p value |
| **CSF** | 0.892 (-17.400-19.184) | 0.924 | 0.326 (-2.798-3.450) | 0.838 | -0.566 (-19.172-18.040) | 0.952 |
| **SI** | -6.355 (-20.883-8.173) | 0.391 | 0.422 (-2.113-2.957) | 0.744 | 6.777 (-8.008-21.562) | 0.369 |
| **SO** | -7.073 (-16.689-2.543) | 0.149 | 0.969 (-0.704-2.643) | 0.256 | 8.042 (-1.742-17.828) | 0.107 |
| **NI** | -1.972 (-16.860-12.915) | 0.795 | -0.421 (-3.076-2.233) | 0.756 | 1.551 (-13.618-16.720) | 0.841 |
| **NO** | -10.438 (-21.641-0.764) | 0.068 | 0.913 (1.096-2.923) | 0.373 | 11.352 (0.065-22.769) | 0.051 |
| **II** | 0.829 (-15.012-16.671) | 0.918 | 0.011 (-2.682-2.706) | 0.993 | -0.817 (-16.925-15.289) | 0.921 |
| **IO** | -8.466 (-18.548-1.615) | 0.100 | 1.236 (-0.481-2.953) | 0.158 | 9.702 (-18.548-1.615) | 0.064 |
| **TI** | 1.156 (-15.447-17.761) | 0.891 | 0.590 (-2.301-3.482) | 0.689 | -0.566 (-17.463-16.330) | 0.948 |
| **TO** | -1.086 (-11.448-9.274) | 0.837 | 0.780 (-1.016-2.577) | 0.394 | 1.867 (-8.674-12.410) | 0.728 |

| Cholesterol | Coefficient (95% Confidence Interval) | p value | Coefficient (95% Confidence Interval) | p value | Coefficient (95% Confidence Interval) | p value |
|---|---|---|---|---|---|---|
| **CSF** | 0.066 (-0.055-0.187) | 0.285 | 0.051 (-0.091-0.195) | 0.480 | -0.014 (-0.195-0.166) | 0.876 |
| **SI** | -0.061 (-0.159-0.035) | 0.215 | 0.021 (-0.096-0.139) | 0.719 | 0.083 (-0.063-0.230) | 0.267 |
| **SO** | -0.050 (-0.116-0.015) | 0.131 | -0.037 (-0.116-0.041) | 0.352 | 0.013 (-0.085-0.111) | 0.797 |
| **NI** | -0.049 (-0.148-0.049) | 0.329 | 0.051 (-0.068-0.170) | 0.400 | 0.100 (-0.048-0.249) | 0.186 |
| **NO** | -0.070 (-0.147-0.005) | 0.069 | -0.040 (-0.132-0.052) | 0.393 | 0.030 (-0.084-0.146) | 0.604 |
| **II** | -0.042 (-0.148-0.063) | 0.435 | 0.023 (-0.101-0.148) | 0.712 | 0.065 (-0.091-0.222) | 0.413 |
| **IO** | -0.017 (-0.088-0.530) | 0.627 | -0.050 (-0.134-0.032) | 0.234 | -0.033 (-0.138-0.071) | 0.536 |
| **TI** | 0.012 (-0.100-0.125) | 0.829 | 0.014 (-0.121-0.150) | 0.837 | 0.001 (-0.168-0.171) | 0.983 |
| **TO** | -0.032 (-0.102-0.036) | 0.356 | -0.035 (-0.119-0.047) | 0.402 | -0.003 (-0.107-0.101) | 0.954 |
| **BUN/Cr** | Coefficient (95% Confidence Interval) | p value | Coefficient (95% Confidence Interval) | p value | Coefficient (95% Confidence Interval) | p value |
| **CSF** | -0.361 (-1.599-0.876) | 0.567 | 0.485 (-0.402-1.373) | 0.283 | 0.847 (-0.622-2.316) | 0.259 |
| **SI** | 0.136 (-0.880-1.154) | 0.792 | -0.175 (-0.910-0.558) | 0.640 | -0.312 (-1.521-0.896) | 0.613 |
| **SO** | 0.211 (-0.472-0.895) | 0.544 | -0.191 (-0.684-0.301) | 0.446 | -0.403 (-1.216-0.408) | 0.330 |
| **NI** | -0.158 (-1.188-0.871) | 0.763 | -0.173 (-0.910-0.562) | 0.643 | -0.15 (-1.232-1.202) | 0.981 |
| **NO** | 0.448 (-0.338-1.236) | 0.264 | -0.338 (-0.904-0.226) | 0.240 | -0.787 (-1.720-0.144) | 0.098 |
| **II** | 0.059 (-1.023-1.141) | 0.915 | -0.283 (-1.054-0.487) | 0.471 | -0.342 (-1.623-0.938) | 0.915 |
| **IO** | 0.637 (-0.064-1.338) | 0.075 | -0.127 (-0.628-0.372) | 0.616 | -0.765 (-1.595-0.064) | 0.071 |
| **TI** | -0.370 (-1.522-0.780) | 0.528 | -0.188 (-1.019-0.641) | -0.656 | 0.182 (-1.185-1.550) | 0.794 |
| **TO** | 0.286 (-0.426-0.998) | 0.431 | -0.150 (-0.662-0.361) | 0.565 | -0.436 (-1.282-0.409) | 0.312 |
| **Ketone** | Coefficient (95% Confidence Interval) | p value | Coefficient (95% Confidence Interval) | p value | Coefficient (95% Confidence Interval) | p value |
| **CSF** | -10.186 (-22.921-2.547) | 0.117 | 24.951 (-43.682-93.584) | 0.476 | 35.137 (-35.038-105.314) | 0.326 |
| **SI** | -0.373 (-11.118-10.371) | 0.946 | 12.793 (-70.721-45.133) | 0.665 | -12.420 (-71.647-46.807) | 0.681 |
| **SO** | -2.297 (-9.367-4.772) | 0.524 | -27.547 (-65.680-10.586) | 0.157 | -25.249 (-64.240-13.741) | 0.204 |
| **NI** | -3.356 (-14.072-7.359) | 0.539 | -4.055 (-63.947-55.836) | 0.894 | -0.699 (-61.772-60.373) | 0.982 |
| **NO** | -2.631 (-11.150-5.888) | 0.545 | -14.353 (-61. 961- 33.255) | 0.555 | -11.721 (-60.263-36.819) | 0.636 |

| | | | | | | |
|---|---|---|---|---|---|---|
| **II** | -1.530 (-12.809-9.749) | 0.790 | -23.858 (-84.482-36.765) | 0.441 | -22.327 (-84.319-39.663) | 0.480 |
| **IO** | -3.717 (-11.278-3.844) | 0.335 | -9.888 (-50.533-30.755) | 0.633 | -6.171 (-47.731-35.388) | 0.771 |
| **TI** | -2.881 (-14.927-9.164) | 0.639 | -24.858 (-89.826-10.110) | 0.453 | -21.976 (-88.405-44.453) | 0.517 |
| **TO** | -1.480 (-9.938-5.977) | 0.697 | -22.536 (-62.796-17.723) | 0.273 | -21.056 (-62.224-20.111) | 0.316 |
| **RAGE** | Coefficient (95% Confidence Interval) | p value | Coefficient (95% Confidence Interval) | p value | Coefficient (95% Confidence Interval) | p value |
| **CSF** | 1.003 (-0.571-2.579) | 0.212 | -0.055 (-0.584-0.472) | 0.836 | -1.059 (-2.729-0.609) | 0.213 |
| **SI** | -0.349 (-1.627-0.928) | 0.592 | -0.000 (-0.434-0.434) | 1.000 | 0.349 (-1.007-1.705) | 0.614 |
| **SO** | -0.764 (-1.576-0.046) | 0.065 | -0.214 (-0.489-0.060) | 0.126 | 0.550 (-0.310-1.410) | 0.210 |
| **NI** | -0.382 (-1.671-0.906) | 0.561 | 0.029 (-0.406-0.465) | 0.894 | 0.412 (-0.953-1.778) | 0.554 |
| **NO** | -1.127 (-2.082-0.171) | **0.021*** | -0.047 (-0.372-0.276) | 0.772 | 1.079 (0.066-2.092) | **0.037*** |
| **II** | -0.512 (-1.873-0.849) | 0.461 | -0.030 (-0.486-0.425) | 0.896 | 0.481 (-0.960-1.924) | 0.513 |
| **IO** | -0.573 (-1.479-0.332) | 0.214 | -0.002 (-0.306-0.301) | 0.988 | 0.571 (-0.388-1.531) | 0.243 |
| **TI** | 0.094 (-1.359-1.549) | 0.898 | -0.067 (-0.561-0.425) | 0.787 | -0.162 (-1.702-1.380) | 0.836 |
| **TO** | -0.880 (-1.741- -0.019) | **0.045*** | -0.132 (-0.423-0.157) | 0.369 | 0.747 (-0.165-1.660) | 0.109 |
| **NEFA** | Coefficient (95% Confidence Interval) | p value | Coefficient (95% Confidence Interval) | p value | Coefficient (95% Confidence Interval) | p value |
| **CSF** | -13.725 (-36.185-8.735) | 0.231 | -3.571 (-17.602-10.459) | 0.618 | 10.153 (-16.469-36.776) | 0.455 |
| **SI** | -11.867 (-29.896-6.161) | 0.197 | -5.603 (-16.922-5.715) | 0.332 | 6.264 (-15.122-27.651) | 0.197 |
| **SO** | -9.572 (-21.833-2.688) | 0.126 | -1.030 (-8.721-6.660) | 0.793 | 8.541 (-6.002-23.085) | 0.250 |
| **NI** | -17.508 (-35.023-0.007) | 0.050 | -6.136 (-17.095-4.822) | 0.272 | 11.371 (-9.349-32.092) | 0.282 |
| **NO** | -15.707 (-29.677--1.738) | **0.028*** | -1.834 (-10.613-6.945) | 0.682 | 13.873 (-2.663-30.411) | 0.100 |
| **II** | -22.627 (-40.895--4.359) | 0.015 | -4.726 (-16.099-6.646) | 0.415 | 17.900 (-3.728-39.529) | 0.105 |
| **IO** | -10.319 (-23.260-2.622) | 0.118 | 0.633 (-7.446-8.712) | 0.878 | 10.952 (-4.377-26.282) | 0.161 |
| **TI** | -16.824 (-37.088) | 0.104 | -5.282 (-17.991-7.425) | 0.415 | 11.541 (-12.494-35.577) | 0.347 |
| **TO** | -12.846 (-25.434--0.258) | 0.045 | -0.357 (-8.233-7.519) | 0.929 | 12.489 (-2.437-27.417) | 0.101 |

Values reported as regression coefficient (95% confidence interval).
Asterisk indicates statistical significance.
Abbreviations: GCL-IPL, ganglion cell layer-inner plexiform layer; SI, superior inner; SO, superior outer; NI, nasal inner; NO, nasal outer; II, inferior inner; IO, inferior outer; TI, temporal inner; TO, temporal outer; SBP, systolic blood pressure; HbA1c, glycated hemoglobin; BUN/Cr, blood urea nitrogen/creatinine; RAGE, receptor for advanced glycation end products; NEFA, non-esterified fatty acids
